# Combined genomic and phenotypic classification of inherited and acquired genetic disease with long-read sequencing

**DOI:** 10.1101/2025.05.08.25327265

**Authors:** Yves Fontaine, Shreegowri Aradhya, Shannon Ji, Etienne Masle-Farquhar, Isabelle Bosi, Julia Forkgen, Zhen Qiao, Igor Stevanovski, Andre L.M. Reis, Melissa Rapadas, Daniel Suan, Peter Hsu, Brynn Wainstein, Alex W. Hewitt, Gemma A. Figtree, Ira W. Deveson, Owen M. Siggs

## Abstract

Current long read sequencing (LRS) platforms allow the simultaneous detection of both genetic variation and epigenetic modification, yet in most cases only genetic variation is utilised. Here we demonstrate the additional potential utility of methylation-based cell type deconvolution and outlier detection, using LRS data from two different platforms. This approach could reliably estimate the proportions of the most abundant cellular constituents of human blood and peripheral blood mononuclear cells, using either primary samples or synthetic mixtures of data from purified cells. Using samples from patients with a hematological malignancy (B cell chronic lymphocytic leukemia) or immunodeficiency (X-linked agammaglobulinemia), LRS resolved both the disease-associated variant and primary cellular phenotype in a single assay. The LRS approach yielded similar cell proportion estimates to orthogonal scRNAseq data generated on the same samples. LRS-derived methylation data therefore represents an incidental source of phenotypic data, with potential future utility in genomic discovery in population-scale biobanks, and in the investigation of inherited and acquired genetic disease.

## Introduction

Short read WGS produces read lengths on the order of 150bp, although current and emerging long read sequencing (LRS) technologies allow the sequencing of reads which are orders of magnitude longer. This increase in read length has some innate advantages over short read WGS including resolution of highly repetitive regions of the genome, large structural variants, and long-range phasing of haplotypes (Logsdon et al. 2020). Though their approaches may differ, current LRS technologies operate by parsing a time series of observations taken directly from long nucleic acid molecules into the nucleotide sequence. One underappreciated advantage of this approach is the ability to read not just the nucleotides of a DNA molecule, but also any epigenetic modifications, as these modifications to the chemical structure of the molecule cause detectable changes in the time series data. Because native genomic DNA molecules are generally sequenced, information of the epigenetic state of cells or tissues from which the material was extracted can be obtained as a byproduct of LRS with no additional assay required, and so presents an opportunity to leverage this data at no additional reagent cost.

Within an individual the genome is largely constant, yet individual cells display an extreme variety of distinct phenotypes, and perform a vast array of functions that are kept unique between the different phenotypes. Epigenetic modifications are responsible for much of this diversity, given the relatively constant genome, and as such epigenetic data can provide important insight into the cell states present in an individual. This information already has significant clinical utility, for example in the diagnosis of neurodevelopmental diseases with characteristic epigenetic signatures (Sadikovic et al. 2021), or for the determination of the cellular origin of cell-free DNA to detect the presence of cancer (Lo et al. 2021). Until recently the epigenetic state of DNA has typically been probed with methylation-sensitive micro-arrays (e.g. Illumina EPIC array) or bisulfite sequencing (Frommer et al. 1992)(Tang 2023).

Here we apply LRS to test the utility of combining genomic and epigenomic information into a single assay. We present a set of epigenetic deconvolution and outlier analyses using LRS data from purified cells, healthy controls, and patients with disorders of B cell development, leveraging LRS to reveal both the genotypic and phenotypic information from the same assay.

## Results

### Robust prediction of leukocyte populations from purified cells and synthetically mixed data

As a proof of concept, we first performed our LRS-derived epigenetic analysis on a dataset consisting of purified leukocyte cells, which we could bioinformatically recombine to create synthetic samples with exact, known compositions (Figure 1A). The six cell types chosen for our analysis were the six most populous cell types of whole blood: Neutrophils, Monocytes, CD8 T Cells, CD4 T Cells, B Cells, and NK Cells, with extracted DNA sequenced on an ONT PromethION instrument. On the resulting dataset we performed methylation-based deconvolution. This works by inferring the proportional cell makeup of a sample based on the frequency of methylation at each genomic locus, given the characteristic epigenetic profiles of each cell type (Moss et al. 2018). This could reliably predict the population of origin for all sorted samples (Figure 1B, Supplementary Table 1).

**Figure 1.**
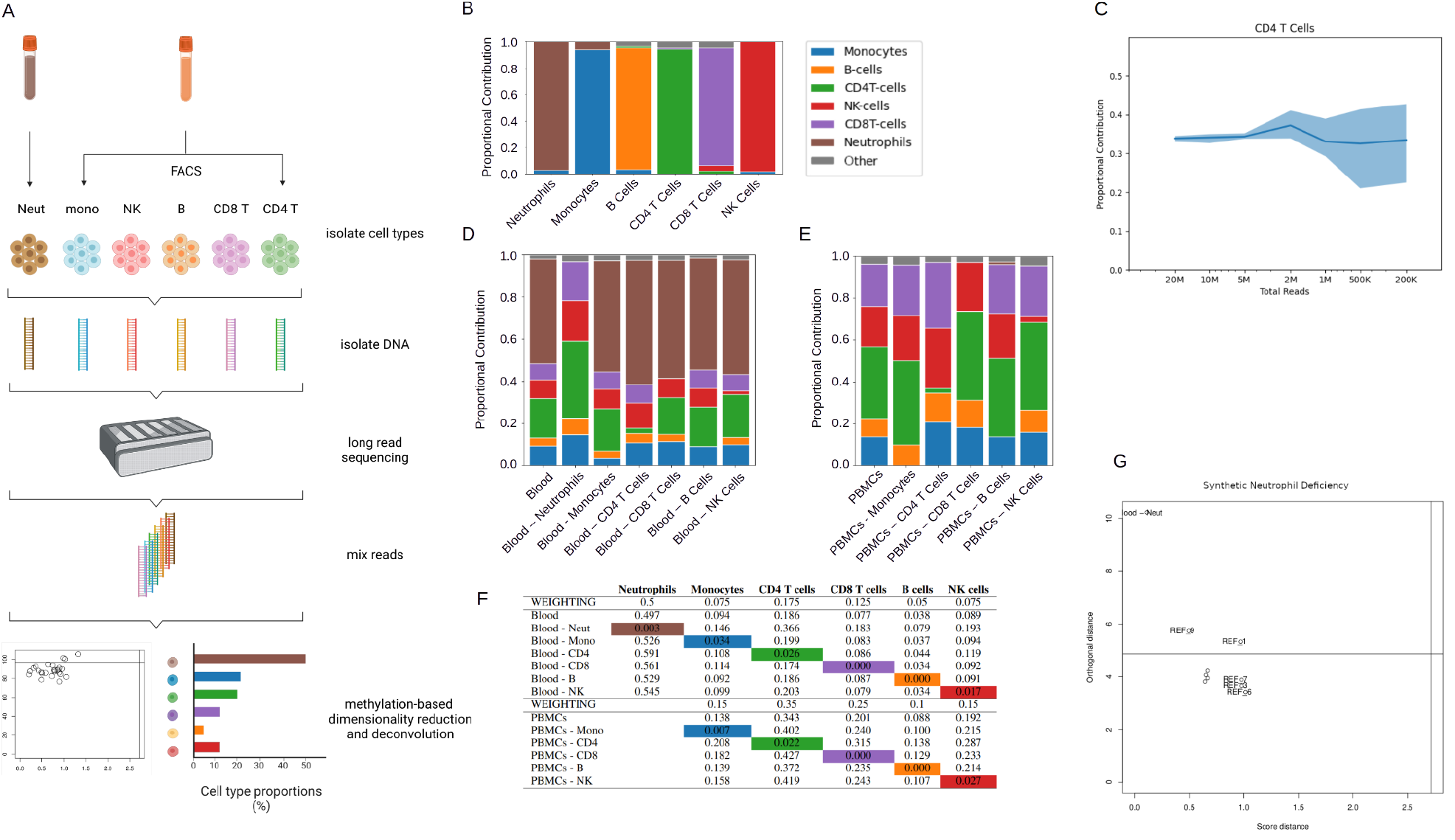
Epigenetic Analysis of Purified Leukocyte Populations and their Synthetic Mixtures. A) Overview of the sorting, sequencing, and synthetic mixing approach. Briefly, purified leukocyte populations were sequenced, and were deconvolved in isolation, or after synthetically combining reads from multiple populations. B) Stacked bar chart showing the results of cell type deconvolution on each isolated leukocyte population alone. C) Graph showing the deconvolution predicted contributions of CD4 T cells to each synthetic PBMC sample at varying total read counts with constant sampling weighting. The shaded region represents the range of predicted values across 3 repeats, while the solid line is the average predicted value. D-E) Stacked bar chart showing the results of cell type deconvolution on the synthetic blood (D), and synthetic PBMC (E) lineage specific deficiency samples. F) Table showing the weightings with which the synthetic deficiency samples were constructed, and the predicted proportions of their deconvolution with respect to the 6 (for whole blood) or 5 (for PBMC) most abundant cell types. G) Results of the outlier detection approach as applied to the synthetic neutrophil deficiency sample alongside a whole blood reference cohort. The vertical and horizontal lines represent an algorithmically determined outlier threshold (Todorov et Filzmoser 2009).

In order to test the reliability of deconvolution from more complex cell mixtures, we then combined LRS data from these purified leukocyte populations to simulate samples of PBMCs (Kleiveland 2015). The weightings used to sample reads from each purified population were set to their typical occurrence in PBMCs while the total number of reads sampled was changed in order to find the variance in deconvolution predictions at different read depths (Figure 1C, Supplementary Figure 1). Following this we combined the LRS data into a series of synthetic whole blood and PBMC samples to simulate lineage-specific leukocyte deficiencies through a “leave one out” approach (Figure 1D-F). Here, the weightings used to sample reads were initially set to their typical occurrence in blood or PBMCs, and were zeroed in the case of each simulated deficiency.

Furthermore, we applied an outlier detection approach previously used in the context of whole-transcriptome RNA sequencing (RNAseq) data (Chen et al. 2020), comparing each whole blood synthetic sample deficient in a given cell type to data from 9 otherwise healthy individuals (Figure 1G, Supplementary Figure 2). In each synthetic deficiency, the methylation-based deconvolution predicted the absence or near absence of the targeted cell type, while the outlier detection approach identified the neutrophil deficiency, but lacked sensitivity for the other deficiencies.

**Figure 2.**
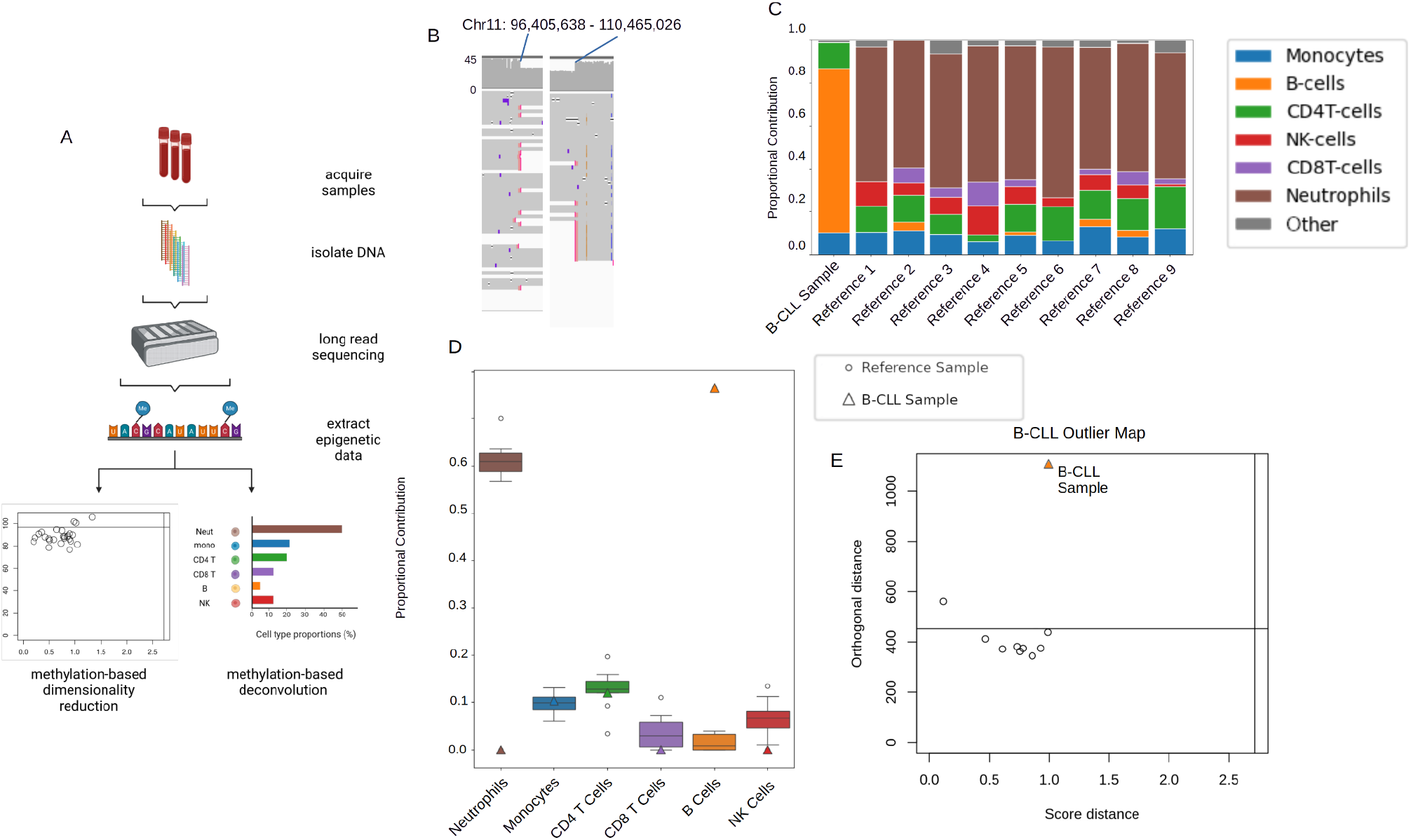
Simultaneous detection of a large somatic deletion, and B cell expansion, in a patient with B-CLL. A) Schematic illustration of the generation and processing of blood-derived LRS data. B) The terminal breakpoints of a chromosome 11 deletion (hg38, chr11: 96,405,638-110,465,026) detected in DNA from a patient with a history of B-CLL,as visualised in IGV (Robinson et al. 2023). The upper track shows the read coverage depth while the lower track shows individual reads. The red marks on the lower track represent reads that are mapped to multiple locations, and they extend over the deletion boundary. C) Stacked bar chart showing the results of cell type deconvolution performed on blood-derived DNA from 10 individuals, including one with a history of B-CLL. D) Box plot showing the deconvolution-derived percentage of each cell type in each sample, as grouped by cell type. Boxes represent the IQR of the data, while whiskers show the data range, discounting outliers as determined by the IQR. Data points from the B-CLL sample are represented with coloured triangles. E) Results of the outlier detection approach. Each circle represents a sample, and the B-CLL sample is represented with an orange triangle. The horizontal line represents an algorithmically determined outlier threshold (Todorov et Filzmoser 2009).

### Simultaneous detection of a large somatic deletion, and B cell expansion, in a patient with chronic lymphocytic leukemia

We next proceeded to test our approach on ONT LRS data from a series of 10 whole blood-derived DNA samples (Supplementary Table 4), one of which was collected from a patient with B cell chronic lymphocytic leukaemia (B-CLL) (Figure 2A). Genome-wide CNV calling detected a large chromosome 11 deletion (normalised read depth 0.744501) in the B-CLL patient (Figure 2B) but none of the other 9 samples, spanning the well-known *ATM* locus (hg38 chr11: 108,222,804-108,369,102) which is recurrently mutated in hematologic malignancies including B-CLL (Schaffner et al. 1999). The same deletion was also detected using structural variant callers in genotyping array data, and short read whole genome sequencing data, with cell frequencies of 0.5312, and 0.5152 respectively (Loh et al. 2018)(Qiao et al., in preparation). Methylation-based cell type deconvolution revealed predicted cell type proportions that were consistent with flow-cytometric reports (Kleiveland 2015), with the exception of a significant overrepresentation of B cells in the B-CLL sample (76.5% compared to a mean of 1.34% in the other 9 samples) (Figure 2C,D, Supplementary Table 2). The same sample was also a clear outlier when applying the outlier detection approach described previously (Figure 2E).

### Simultaneous detection of a missense variant and B cell Deficiency in a patient with X-linked agammaglobulinemia

Finally, we tested the utility of our approach in the context of monogenic leukocyte deficiencies. Such patients may undergo whole genome sequencing, given their genomic and phenotypic heterogeneity (Bousfiha et al. 2022), but their genetic diagnoses are not always resolved by srWGS. One such disease is X-linked agammaglobulinemia (XLA): a disorder most commonly affecting males, and characterised by a deficiency of B cells and the immunoglobulin molecules they secrete.

We generated PBMC-derived LRS data from a single XLA patient on the PacBio Revio instrument, and compared it to data derived from 25 reference samples prepared under the same conditions (Supplementary Table 4). LRS successfully revealed a known pathogenic missense variant in *BTK* (p.Arg520Gln, absent from gnomAD v4.1.1, ClinVar accession number VCV000011378.8), known to be responsible for the patient’s presentation. The patient’s percentage of CD19+ B cells on clinical flow cytometry was <1% (<0.05 x10^9^/L, reference range 0.2 − 2.1). Consistent with this, methylation-based deconvolution of the LRS data revealed a predicted B cell proportion of 0.00% in the XLA patient sample (Figure 3B-C, Supplementary Table 3), although due to high variance in the reference data this was not statistically significant (B cell reference mean 7.97%, SD 4.77%), nor was it detected through the outlier detection method (Figure 3D). Three samples in the reference cohort however were identified as potential outliers. As an orthogonal measure, we compared these values predicted for the reference cohort to estimates from paired single cell RNAseq data (Figure 3E), which showed a general agreement in proportional estimates, with a tendency for LRS-derived proportions to be higher than those from scRNAseq for CD4 T cells and monocytes, while the reverse was found for CD8 T cells and B cells, likely demonstrating the cell type biases specific to each platform (Newman et al. 2019).

**Figure 3.**
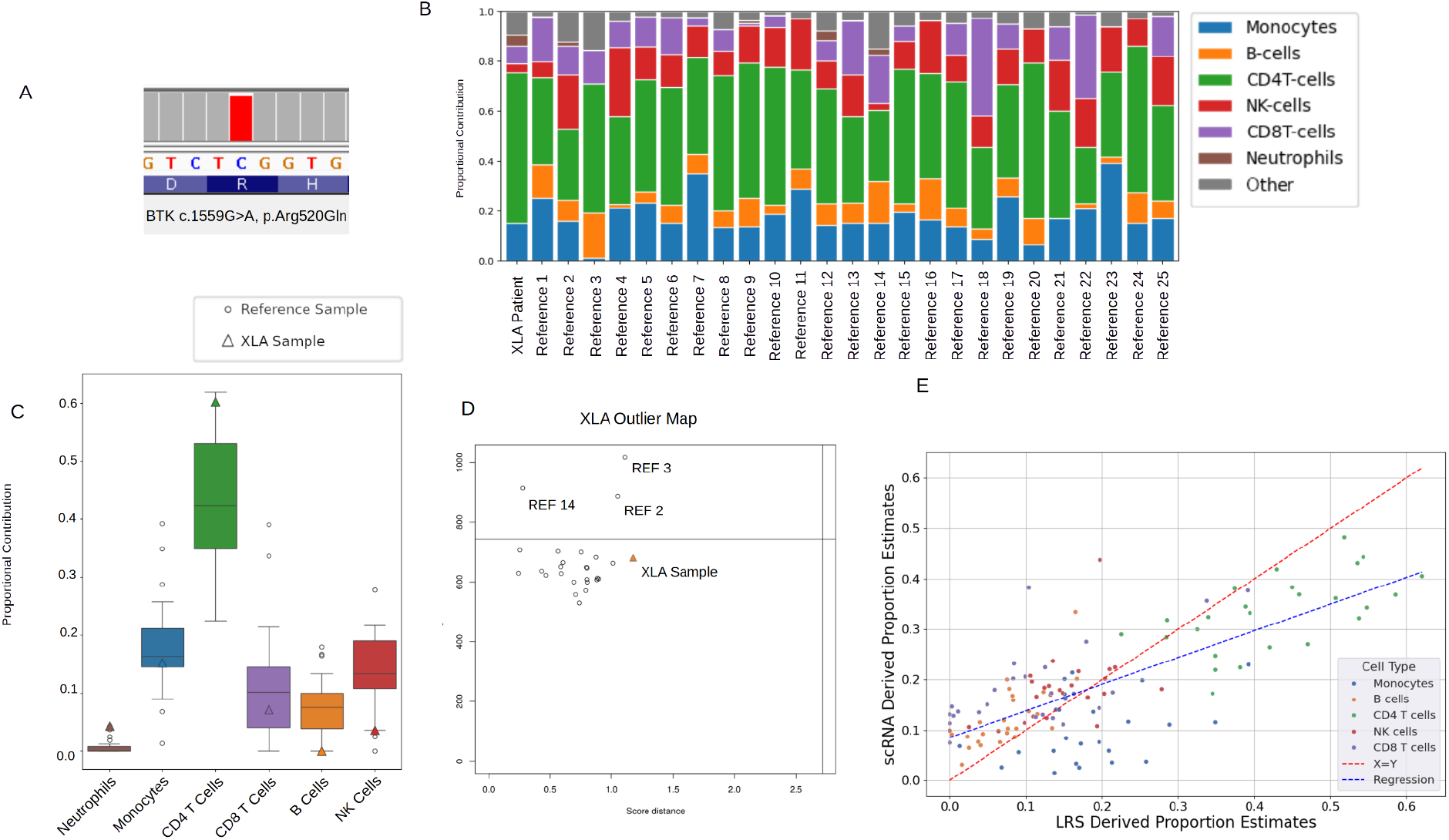
Simultaneous detection of a *BTK* missense variant, and B cell deficiency, in a patient with XLA. A) The c.1559G>A p.Arg520Gln variant in *BTK* responsible for XLA, as visualised in IGV. B) Stacked bar chart showing the results of PBMC cell type deconvolutions performed on the XLA patient and 25 reference samples. C) A box chart showing the percentage contribution of each cell type to its sample, as grouped by cell type and predicted from the deconvolution. Boxes represent the IQR of the data, while whiskers show the data range, discounting outliers as determined by the IQR. The data points representing the XLA sample are shown in coloured triangles. D) Results of the outlier detection approach as applied to the XLA sample and PBMC reference dataset. Each circle represents a sample, with the XLA sample and several reference samples labelled. The horizontal line represents an algorithmically determined outlier threshold (Todorov et Filzmoser 2009). E) Difference in the predicted proportional contributions to each sample of a given cell type, as predicted from LRS methylation data (X-axis), and from paired single cell RNA sequencing data (Y-axis). Different cell types are represented by different colours. The red diagonal line represents the identity function X=Y, with the regression line in blue.

## Discussion

This work presents a series of three datasets derived from two LRS platforms on which we have performed both genetic and epigenetic analyses, including methylation-based cell type deconvolution and outlier detection based approaches. Deconvolution reliably predicted the proportional cell type composition of each sample, while an outlier detection method was less sensitive at identifying phenotypic outliers against a small reference cohort (9-25 samples). In the isolated leukocyte populations the deconvolution predicted each cell type to be almost entirely composed of that cell type, wherein the error potentially arises from sorting impurities and the imprecise nature of epigenetic deconvolution. Given that our deconvolution model was a non-negative least squares regression model (Moss et al. 2018), some predictive error was to be expected.

In synthetically recombined samples, most cell type proportions predicted by deconvolution were approximately equal to the proportions with which these samples were constructed. For example, neutrophils, CD8 T cells, and B cells were uniformly absent from deconvolution outputs when absent from the synthetic mixtures. Some notable exceptions were CD4 T cells and NK cells, which in spite of an absence of reads in the synthetic mixtures, were still predicted in small proportions in the deconvolution outputs. It is possible that these cell types simply have a less distinct epigenetic signature, resulting in greater error when applying the regression model.

In two different clinical extremes (a pathogenic leukocyte expansion, and a pathogenic leukocyte deficiency), methylation-based deconvolution reliably predicted both from LRS data, although the dramatic expansion of B cells in the context of B-CLL was more obvious than the deficiency observed in XLA, particularly in the context of variable B cell proportions amongst reference samples. Similarly, an outlier detection-based approach to detecting anomalous samples was less sensitive. This approach only detected outliers where samples differed significantly from the reference cohort (e.g. a lack of neutrophils, or large B cell expansion), but was not sensitive enough to detect smaller differences (e.g. a lack of a rarer cell subset such as B cells).

### Limitations and Future Work

In both our deconvolution and outlier approach, the sensitivity appears to improve where more reads are derived from the cell population of interest. This is to be expected, as a more common cell type will contribute more DNA to the sequenced sample, and therefore the true proportion is predicted with more reliability. For example, the key difference between the XLA sample and the 25 PBMC reference samples was an absence of B cells in the XLA sample. However, in the reference samples the average B cell proportion was found to be 7.97% with a standard deviation of 4.77%. Even though the XLA sample was the only sample with a predicted B cell proportion of 0%, this was not a statistically significant deviation from the reference cohort distribution. As the scale of population reference data increases, this is likely to become less of an issue.

Sensitivity is also influenced by sequencing depth, which at an average of ~36X for ONT sequenced samples, and ~30X for PacBio sequenced samples (Supplementary Table 7), means that ~3 reads are derived from B cells in PBMCs, and even fewer in whole blood. Deeper sequencing coverage, either genome-wide, or at specific differentially methylated loci, is one way to overcome this. This lower sensitivity combined with the cost of LRS limits the value of our approach in the clinical setting at the individual level, compared with the current standard of clinical flow cytometry, though future advancements may change this.

As LRS becomes more commonplace, there are increasing opportunities to use methylation data to infer phenotype. The outlier detection approach used here was selected as it had been employed previously on RNAseq data: a similarly high dimensional, low sample count context in the biological domain, however this approach may benefit from a methylation-specific approach. Similarly, the increasing volume of public LRS datasets will create larger reference cohorts, and increase the power with which phenotypic outliers can be detected.

Another consideration is the selection of differentially methylated loci. We investigated the performance differences in using all >27 million methylated loci detected by LRS; the subset of ~450,000-850,000 loci available to bisulfite arrays; and the smallest subset of ~8000 differentially methylated sites used by the *methatlas* deconvolution package. We found that smaller subsets of loci had improved performance over the larger sets (Supplementary Figure 3), perhaps pertaining to the increased signal to noise ratio and greater information density. In each dataset, we used the largest number of loci which still produced a meaningful result, with the ~8000 *methatlas* loci in the synthetic and the XLA data, while the 27.8 million sites, without curation, were sufficient for the B-CLL data. Nonetheless, it is likely that improved performance could be gained from further refining the loci used in this approach.

Additionally, with respect to cell type deconvolution, our work has focused on the use of only one specific tool (*methatlas*), whose deconvolution matrix was derived from the limited number of sites present in methylation array data. Epigenetic deconvolution allows for greater cell type resolution over other recently reported sequencing-based approaches (Bentham et al. 2025; Poisner et al. 2024), and its accuracy will only improve as new models and reference datasets are developed (Loyfer et al. 2023)(Salas et al. 2022), including the potential for LRS-specific tools which leverage the >27 million sites assayed by long read derived data.

## Conclusions

Our work demonstrates that methylation-based deconvolution of LRS data is able to reliably predict the cellular composition of a sample. We demonstrated its application in two clinical scenarios (hematological malignancy, and immunodeficiency), while simultaneously revealing the underlying genetic cause. Although blood and PBMCs were the reference samples used here, the same approach could be used for any DNA sample with an appropriate deconvolution reference (e.g. tumour biopsies, saliva etc). Given the cost of LRS at the individual scale, however, in the nearer term this approach is likely to be of significant value in the context of population-scale biobanks, where methylation-based deconvolution can be applied across large LRS datasets to discover new associations between genetic variation and cell-type abundance (Rumker et al. 2024). As LRS technology matures, methylation data will become increasingly abundant and accessible for orthogonal analyses. Increasingly large reference datasets, deeper and more accurate coverage, and more sophisticated methods are all emerging: as such we can expect the utility of LRS-derived methylation data to only continue to improve.

## Data Availability

All data referred to in the present study will be available at the time of publication if allowable by their respective ethics protocols

## Funding and Acknowledgements

This research was supported by the Snow Medical Research Foundation (Grant No. PF2019-040). The funders were not involved in the design of the study, collection, analysis, and interpretation of the data, the writing of this report, or the decision to submit the article for publication. The authors express their sincere appreciation to the Snow Medical Research Foundation for their generous support of this work.

## Methods

### Cohorts and samples

CD4+ T cells, CD8+ T cells, Monocytes, B cells, and NK cells were purified from blood of healthy donors (Western Sydney Local Health District Human Research Ethics Committee HREC 17/5449). Magnetically-enriched neutrophils were sourced from Stem Cell Technologies. The demographic breakdown of each sample donor is available in Supplementary Table 4. Where possible, the demographics of the neutrophil donors were matched to the PBMC donors.

Blood-derived long-read data was generated from a subset of samples from the Tasmanian Ophthalmic Biobank (TOB)(Yazar et al. 2022) (University of Tasmania HREC 2020/ETH02479). This dataset included a total of 10 individuals of European ancestry sampled from the general population, including one individual with B cell chronic lymphocytic leukaemia (B-CLL). The cohort demographics are described by Yazar et al., with a demographic breakdown of the samples provided in Table Supplementary Table 4. In addition to the processing described herein, this cohort was analysed by genotyping array, and srWGS methods for structural variants as previously described (Loh et al. 2018; Tanudisastro et al. 2024).

PBMC-derived long read data was generated from a subset of samples from the BioHeart cohort (Northern Sydney Local Health District HREC/17/HAWKE/343) (Tanudisastro et al. 2024; Kott et al. 2019). This dataset included 25 individuals of European ancestry with diagnosed or suspected cardiac disorders, and is described in greater detail by Tanudisastro et al. Additionally, PBMC-derived data was also generated from a patient diagnosed with XLA (Sydney Children’s Hospital Network HREC/18/SCHN/140).

### Fluorescence-activated cell sorting of leukocytes

To purify the CD4+ T cells, CD8+ T cells, Monocytes, B cells, and NK cells, fluorescence-activated cell sorting (FACS) was used. PBMC samples were thawed and washed with PBS 2% FCS and counted with the Countess 3 Automated Cell Counter. Cells were stained using the fluorophore-conjugated monoclonal antibody mixture described in Supplementary Table 5, for 25 minutes under low light conditions. Cells were then washed with PBS 2.5% FCS, and centrifuged at 1500 rpm for 5 minutes at 4C, followed by resuspension in PBS 5% FCS for sorting using BD FACSAria 2 and FACSAria 2B machines. Cell purity was additionally checked on the FACSAria 2 machine and is reported in Supplementary Table 6. The reported NK cell purity is likely an underestimate due to a technical artifact. Gates used for sorting and purity checks are provided in Supplementary Figure 4. Sorted cell populations were centrifuged at 2,000g for 4 minutes at 4C, the supernatant discarded and cell pellets snap frozen at −80C for subsequent DNA extractions.

Vials of purified neutrophils (>10^6^ cells) at a purity of ≥95% were isolated from peripheral blood by negative immunomagnetic separation (Stem Cell Technologies), and preserved in CryoStor CS10 medium with ACDA as an anticoagulant. Vials were thawed and washed with PBS, 2% FCS solution and the cells were pelleted at 1500 rpm for 5 minutes, 4C, before resuspension in PBS, 5% FCS, for counting. Cells were then counted with a Countess 3 Automated Cell Counter, and 300 uL from each sample were combined into a sample totaling approximately 3 million cells.

These were re-pelleted at 500G for 5 minutes, at 4C.

### DNA extraction and quality control

DNA extraction from the purified leukocyte populations, BioHeart cohort, and XLA patient was performed using the ‘Nanobind HMW DNA from cultured cells’ kit (102-301-900, PacBio) following manufacturer instructions. Briefly, cells were lysed and treated with proteinase and RNAse. The RNAse activity was then neutralised by a change of buffer. DNA was then precipitated onto a magnetic nanobind disc and eluted.

For the TOB cohort, DNA extraction was extracted from whole blood using ‘DNeasy Blood and Tissue’ kits (69504, Qiagen) following manufacturer instructions. Briefly, cells were lysed and DNA was isolated with a silica-based spin column, followed by elution.

All DNA was quality checked for concentration using Qubit (Thermo Fisher Scientific), purity/quality using a Nanodrop (Thermo Fisher Scientific), and for fragment size distribution using FemtoPulse (FemtoPulse genomic DNA 165kb analysis kit, Agilent). Ideal Qubit readings required averaging of 3 readings taken from top, middle and bottom of 1.5mL DNA LoBind tubes. Ideal quality check (QC) readings are as follows: Qubit (>15ng/µL for >1 µg DNA output), Nanodrop (260/280 reading > 1.8, and 260/230 reading > 2.00) and Femto pulse (single high peak). Extracted DNA that met all QC checks was stored at −20C.

### ONT sequencing

The TOB, and purified leukocyte populations were sequenced on an ONT PromethION as previously described (Reis et al. 2023). DNA quantity was measured using a Qubit (Thermo Fisher Scientific), purity on a NanoDrop (Thermo Fisher Scientific) and fragment-size distribution on a TapeStation (Agilent). Prior to ONT library preparations, DNA was sheared to ~15–20 kb fragment size using Covaris G-tubes at 4200rpm. No shearing was performed on samples where the starting fragment distribution peaked at or below ~25 kb. Sequencing libraries were prepared from ~1–2ug of DNA, using native library preparation kits (either SQK-LSK110 or SQK-LSK114), according to the manufacturer’s instructions. Each library was loaded onto a PromethION flow cell (R9.4.1 for SQK-LSK110 libraries, R10.4.1 for SQK-LSK114 libraries) and sequenced on an ONT PromethION P48 device. Samples were run for a maximum duration of 72 h, with 1–3 nuclease flushes and reloads performed during the run, where necessary to maximize sequencing yield Raw ONT sequencing data were converted from FAST5 to the more compact BLOW5 format in real-time on the PromethION during each sequencing run using slow5tools (v.0.3.0). BLOW5 data were transferred to the Australian National Computational Infrastructure (NCI) high-performance computing environment before further processing. Data were base-called with Guppy (v.6.0.1), using the Buttery-eel wrapper for BLOW5 input, with the high-accuracy model and reads with mean quality <7 were excluded from further analysis. Methylation calling was performed with f5c.

The resulting unmapped bam files were aligned to the hg38 human reference genome using Minimap2, and, as this process does not preserve the methylation tags, mapped reads were re-tagged with the methylation tags from the unmapped bam file. The Illumina Bead Chip loci were then lifted to hg38, and used to produce a csv of methylation frequency per Bead Chip index. This was done manually as, at the time, WGBStools did not yet have the capability to extract the frequency per Illumina index for an hg38 mapped sample directly.

### HiFi sequencing and data processing

DNA samples were homogenized using a Diagenode Megaruptor with the 3 DNAFluid+ Kit (E07020001, Diagenode) under the following conditions: volume 150 µL, speed 40, and concentration 50 ng/µL. Subsequently, 3 µg of material was diluted in low TE to a final volume of 130 µL. Shearing was performed with the Megaruptor shearing kit (E07010003, Diagenode) at speed 30 or 31, aiming for average fragment lengths of 15–24 kb. Clean-up and concentration of the sheared material were conducted using SMRTbell clean-up beads (102-158-300, PacBio), and the DNA was eluted in 47 µL. Average fragment lengths were determined using the Femto Pulse system with the Genomic DNA 165kb Analysis Kit (FP-1002-0275, Agilent). SMRTbell libraries were prepared using the SMRTbell® Prep Kit 3.0 (102-141-700, PacBio) following standard procedures, including unique barcoding of each sample with the SMRTbell Adapter Index Plate 96A (102-009-200, PacBio). Size selection was performed with AMPure PB beads (102-182-500, PacBio) at a 2.9x ratio. Final library sizes were confirmed with the Femto Pulse, and libraries were diluted to below 60 ng/µL before the ABC loading procedure. SMRT library fragment lengths ranged between 12.895 and 26.001 kb. Sequencing was performed using the following PacBio products: Revio sequencing plate (02-587-400), Revio Polymerase kit (102-739-100), and Revio SMRT cell tray (102-202-200). On-plate loading concentration was set to 250 pM, with 30-hour movie times. SMRT libraries that did not yield 90 Gb using one SMRT cell were pooled for additional sequencing in “top-up” runs

### Synthetic mixtures

The creation of our synthetic samples was performed at the read level, and began with the mapped bamfiles of the purified leukocyte populations. Sampling the reads of these bamfiles was performed using samtools’ view function, with the ‘-s’ flag. In creating synthetic PBMC samples to test deconvolution prediction variance at different read depths, total read counts of 20, 10, 5, 2, and 1 million, as well as 500, and 200 thousand were used. In creating each synthetic leukocyte deficiency sample, 20 million total reads were used in the PBMC samples, while 10 million were used in the blood samples so as to not over-sample the neutrophil reads. The weightings with which each purified population is sampled in a given synthetic sample are given in Figure 1 Panel E. The weights used are as presented in the table save for that they are set to zero in samples intended to be deficient in a given population. Note that while these weightings were chosen to be equivalent to percentages in the synthetic healthy whole blood and PBMC samples, they are not percentages. Consequentially, setting the weight of a cell population to zero in a given sample increases the sampling rate of the other populations in proportion with their weightings. For example, in the synthetic blood sample deficient in neutrophils, the weighting for neutrophils was set to zero where it was initially 0.5, which effectively doubled the expected proportions of the remaining cell types. This process produced 13 synthetic samples in total, including 1 healthy whole blood sample, and 6 blood samples deficient in one key cell type, alongside 1 healthy PBMC sample, and 5 PBMC samples deficient in one key cell type. The frequency of methylation per chromosomal locus was extracted with WGBStools (Loyfer et al. 2023) using the bam_to_pat function, which produced pat and beta files. These were additionally converted to the methylation frequency per Illumina Bead Chip Index using the ‘convert’ function of WGBStools to create a mapping between hg38 and Illumina indices followed by manual conversion, though recent updates have allowed this to be performed with the beta_to_450k function directly instead.

### Deconvolution

Deconvolution was performed on our samples in order to predict their proportional cell type composition. This was done using Methatlas (Moss et al. 2018). Methatlas was developed using bisulfite sequencing data, and employs a non-negative least squares regression model to predict cell type compositions. It accepts a CSV with columns of samples, and rows of Illumina Bead Chip Indices, with entries of methylation frequency. Following deconvolution all cell types which were not either Neutrophils, Monocytes, CD4+ T cells, CD8+ T cells, B cells, or NK cells were collapsed into an ‘Other’ category for simplicity.

### Dimensionality reduction

To perform our outlier detection approach, we began with the beta and pat files produced by WGBStools, which contained the methylation frequency per chromosomal locus. These were extracted per-sample using the WGBStools beta_to_bed function. The loci were then filtered for null values, and the intersection of all remaining loci between the samples being analysed was taken to combine samples into a tsv. This typically resulted in approximately 27.8 million loci, with slight variation between cohorts as different loci were filtered out. These loci were then subsetted to the approximate 850,000 or 450,000 loci present in bisulfite sequencing data, and to the approximate 8000 loci used by methatlas, though with imperfect mapping to the hg38 reference genome this was in practice closer to 6800 loci. The following was performed on each subset of the loci, and the largest subset which still produced a meaningful result was retained:

From this process, a matrix with rows representing samples, and columns representing loci was constructed, with entries of methylation frequency. robust PCA with the projection pursuit method, as implemented in the rrcov (Todorov et Filzmoser 2009) package in R, specifically, the ‘PCAGrid’ function, was then performed on this matrix to produce the outlier maps presented herein. This approach was chosen as it has been used previously for high dimensionality, low sample count data in the biological domain, on transcriptomic data (Chen et al. 2020), though it is data type agnostic. The implementation details of this approach are described by (Todorov et Filzmoser 2009). Note that the quadrants are determined algorithmically, and that outliers are expected to appear in the top left quadrant.

